# Effects of New York’s Executive Order on Face Mask Use on COVID-19 Infections and Mortality: A Modeling Study

**DOI:** 10.1101/2020.10.26.20219527

**Authors:** Mingwang Shen, Jian Zu, Christopher K. Fairley, José A. Pagán, Bart Ferket, Bian Liu, Stella S. Yi, Earle Chambers, Guoqiang Li, Yuming Guo, Libin Rong, Yanni Xiao, Guihua Zhuang, Alexis Zebrowski, Brendan G. Carr, Yan Li, Lei Zhang

**Affiliations:** China-Australia Joint Research Center for Infectious Diseases, School of Public Health, Xi’an Jiaotong University Health Science Center, Xi’an, Shaanxi, China; School of Mathematics and Statistics, Xi’an Jiaotong University, Xi’an, Shaanxi, China; School of Electrical Engineering, Xi’an Jiaotong University, Xi’an, Shaanxi, China; Department of Epidemiology and Preventive Medicine, School of Public Health and Preventive Medicine, Monash University, Melbourne, Australia; Department of Mathematics, University of Florida, Gainesville, FL, USA; Department of Population Health Science and Policy, Icahn School of Medicine at Mount Sinai, New York, NY, USA; Department of Obstetrics, Gynecology, and Reproductive Science, Icahn School of Medicine at Mount Sinai, New York, NY, USA; Melbourne Sexual Health Centre, Alfred Health, Melbourne, Australia; Central Clinical School, Faculty of Medicine, Nursing and Health Sciences, Monash University, Melbourne, Australia; Department of Epidemiology and Biostatistics, College of Public Health, Zhengzhou University, Zhengzhou, Henan, China; Department of Public Health Policy and Management, College of Global Public Health, New York University, New York, NY, USA; Leonard Davis Institute of Health Economics, University of Pennsylvania, Philadelphia, PA, USA; Department of Population Health, NYU Grossman School of Medicine, New York, NY, USA; Department of Family and Social Medicine, Albert Einstein College of Medicine, Montefiore Health System, Bronx, NY, USA; Department of Emergency Medicine, Icahn School of Medicine at Mount Sinai, New York, NY, USA

**Keywords:** COVID-19, Mitigation strategy, Non-pharmaceutical intervention

## Abstract

**Background:** New York City (NYC) was the epicenter of the COVID-19 pandemic in the United States. On April 17, 2020, the State of New York implemented an Executive Order that requires all people in New York to wear a face mask or covering in public settings where social distancing cannot be maintained. It is unclear how this Executive Order has affected the spread of COVID-19 in NYC.

**Methods:** A dynamic compartmental model of COVID-19 transmission among NYC residents was developed to assess the effect of the Executive Order on face mask use on infections and deaths due to COVID-19 in NYC. Data on daily and cumulative COVID-19 infections and deaths were obtained from the NYC Department of Health and Mental Hygiene.

**Results:** The Executive Order on face mask use is estimated to avert 99,517 (95% CIs: 72,723-126,312) COVID-19 infections and 7,978 (5,692-10,265) deaths in NYC. If the Executive Order was implemented one week earlier (on April 10), the averted infections and deaths would be 111,475 (81,593-141,356) and 9,017 (6,446-11,589), respectively. If the Executive Order was implemented two weeks earlier (on April 3 when the Centers for Disease Control and Prevention recommended face mask use), the averted infections and deaths would be 128,598 (94,373-162,824) and 10,515 (7,540-13,489), respectively.

**Conclusions:** New York’s Executive Order on face mask use is projected to have significantly reduced the spread of COVID-19 in NYC. Implementing the Executive Order at an earlier date would avert even more COVID-19 infections and deaths.

## INTRODUCTION

The 2019 novel coronavirus (SARS-CoV-2, which causes the disease COVID-19) has been spreading at an alarming rate in the United States (US) and globally.^1,2^ In the US, New York City (NYC) was the epicenter of the pandemic and has the highest mortality rate among all the major cities. As of August 25, 2020, more than 230,000 cases of COVID-19 have been confirmed in NYC, resulting in over 23,000 deaths.^3^ As the largest and most densely populated city in the US, NYC faces incomparable challenges in the social distancing needed to contain the spread of the virus.^4,5^

On April 3, 2020, the US Centers for Disease Control and Prevention (CDC) recommended “wearing cloth face coverings in public settings where other social distancing measures are difficult to maintain (e.g., grocery stores and pharmacies) especially in areas of significant community-based transmission.”^6^ Although face mask use by the general public has been recommended in many countries to reduce COVID-19 transmissions,^7–9^ it was for the first time being recommended in the US. There is growing evidence that face masks can protect the wearer from being infected by respiratory diseases or infecting others.^10–12^ On April 17, 2020, an Executive Order was implemented that requires all residents over age 2 in New York to wear masks or face coverings when they are in public and social distancing is impossible.^13^ However, it is unclear how this Executive Order would affect the spread of COVID-19 in NYC.

This study aims to use a dynamic compartmental model to estimate the effect of the Executive Order on face mask use on COVID-19 infections and deaths. We used COVID-19 data in NYC and the best available evidence to inform model parameters. We estimated the averted numbers of infections and deaths due to the Executive Order on face mask use. We also studied what would happen if the Executive Order was implemented earlier. Findings from this study will provide policymakers with important information on the effect of face mask related policies in NYC and other parts of the US.

## METHODS

### Data sources

We obtained the NYC COVID-19 data from the NYC Department of Health and Mental Hygiene (DOHMH).^3^ Our data included the number of daily and cumulative confirmed cases and deaths from February 29 to June 7, 2020 (**Table S1**). We intentionally chose the last day of evaluation to be June 7, a day before NYC initiated its first phase of reopening, as social distancing is likely to change after June 7. We estimated the disease progression and individual behavioral parameters from the literature (**Table S2**).

### Model formulation and assumptions

We developed a dynamic compartmental model to describe the transmission of COVID-19 in NYC. **Figure 1** shows the structure of our model. Specifically, the NYC population was divided into nine compartments, including susceptible individuals (S), latent individuals (E), asymptomatic infections (A, infected individuals without symptoms), undiagnosed infections with mild/moderate (I_1_) and severe/critical symptoms (I_2_), diagnosed infections with mild/moderate (T_1_) or severe/critical symptoms (T_2_), recovered (R) and deceased (D) cases. Susceptible individuals could be infected by contacts with latent, asymptomatic, and undiagnosed infections with mild/moderate and severe/critical symptoms in public settings (e.g., public transportation, supermarkets, offices, etc.) and households (home or other private settings) with a probability /l (the force of infection) (a detailed description of /l was provided in the **Supplementary Appendix**).

**Figure 1.**
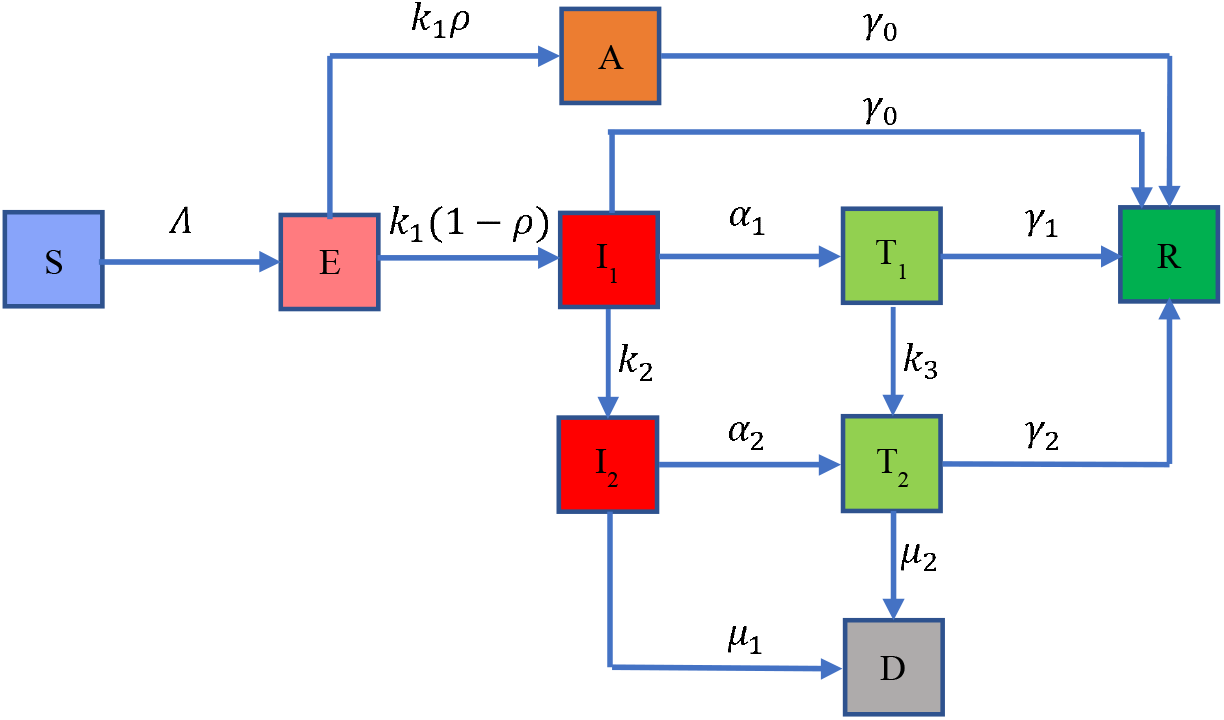
Flow chart of COVID-19 transmission model in New York City. The total population is divided into nine compartments: susceptible individuals (S), latent individuals (E), asymptomatic infections (A), undiagnosed infections with mild/moderate (I_1_) and severe/critical symptoms (I_2_), diagnosed infections with mild/moderate (T_1_) and severe/critical symptoms (T_2_), recovered (R) and deceased (D) cases. The force of infection is denoted as /l, which involves two transmission patterns: public settings (e.g., public transportation, supermarkets, offices, etc.) and households. The model includes three control measures: handwashing, social distancing and face mask use. More details on /l are provided in the **Appendix**. The average incubation period is 1/*k*_1,_ and the probability that an individual is asymptomatic is *ρ*. Infectious individuals with mild/moderate and severe/critical symptoms are diagnosed and treated at the rates *α*_l_and *α*_2_, respectively. We assume these diagnosed individuals are isolated strictly and could not further infect others. Undiagnosed and diagnosed mild/moderate cases progress to the severe/critical stage at the rates *k*_2_ and *k*_3_, respectively. Asymptomatic infections and undiagnosed mild/moderate cases are assumed to recover naturally at the rate *γ*_0_. Diagnosed mild/moderate and severe/critical cases will recover at the rates *γ*_l_ and *γ*_2_, respectively. Undiagnosed and diagnosed severe/critical cases will die due to the disease at the rates *µ*_l_ and *µ*_2_, respectively.

Latent individuals progress to the infectious compartment with mild/moderate symptoms or asymptomatic compartment at a rate *k*_1,_ and the probability that an individual is asymptomatic is *ρ*. Infectious individuals with mild/moderate and severe/critical symptoms are diagnosed and treated at the rates *α*_l_and *α*_2_, respectively. We assumed these diagnosed individuals were isolated strictly and could not further infect others. Undiagnosed and diagnosed mild/moderate cases progress to the severe/critical stage at the rates *k*_2_and *k*_3_, respectively. Asymptomatic infections and undiagnosed mild/moderate cases are assumed to recover naturally at the rate *γ*_O_. Diagnosed mild/moderate and severe/critical cases will recover at the rates *γ*_l_ and *γ*_2_, respectively. Undiagnosed and diagnosed severe/critical cases will die due to the disease at the rates *µ*_l_ and *µ*_2_, respectively.

The CDC recommended face mask use by the general public on April 3, two weeks before the Executive Order on face mask use was implemented on April 17 in New York.^13^ In the baseline analysis, we assumed that the proportion of face mask use by the general public increased from 0% on April 3 to 100% on April 17 when the Executive Order was implemented. While the Executive Order has been enforced to some degree, it is possible that some people may not follow the order. Thus, we varied the proportion of face mask use in the sensitivity analysis to examine the robustness of our results with regards to this parameter. We also assumed that the baseline effectiveness of face mask in preventing COVID-19 infection or infecting others is 85% (95% CI, 66%-93%) based on a recent meta-analysis on the effectiveness of face masks for COVID-19.^12^ Since there are a variety of face masks (from cloth masks to surgical masks), which may increase the uncertainty in face mask effectiveness, we varied the face mask effectiveness from 0 to 100% in our sensitivity analysis (**Figure 3**).

**Figure 2.**
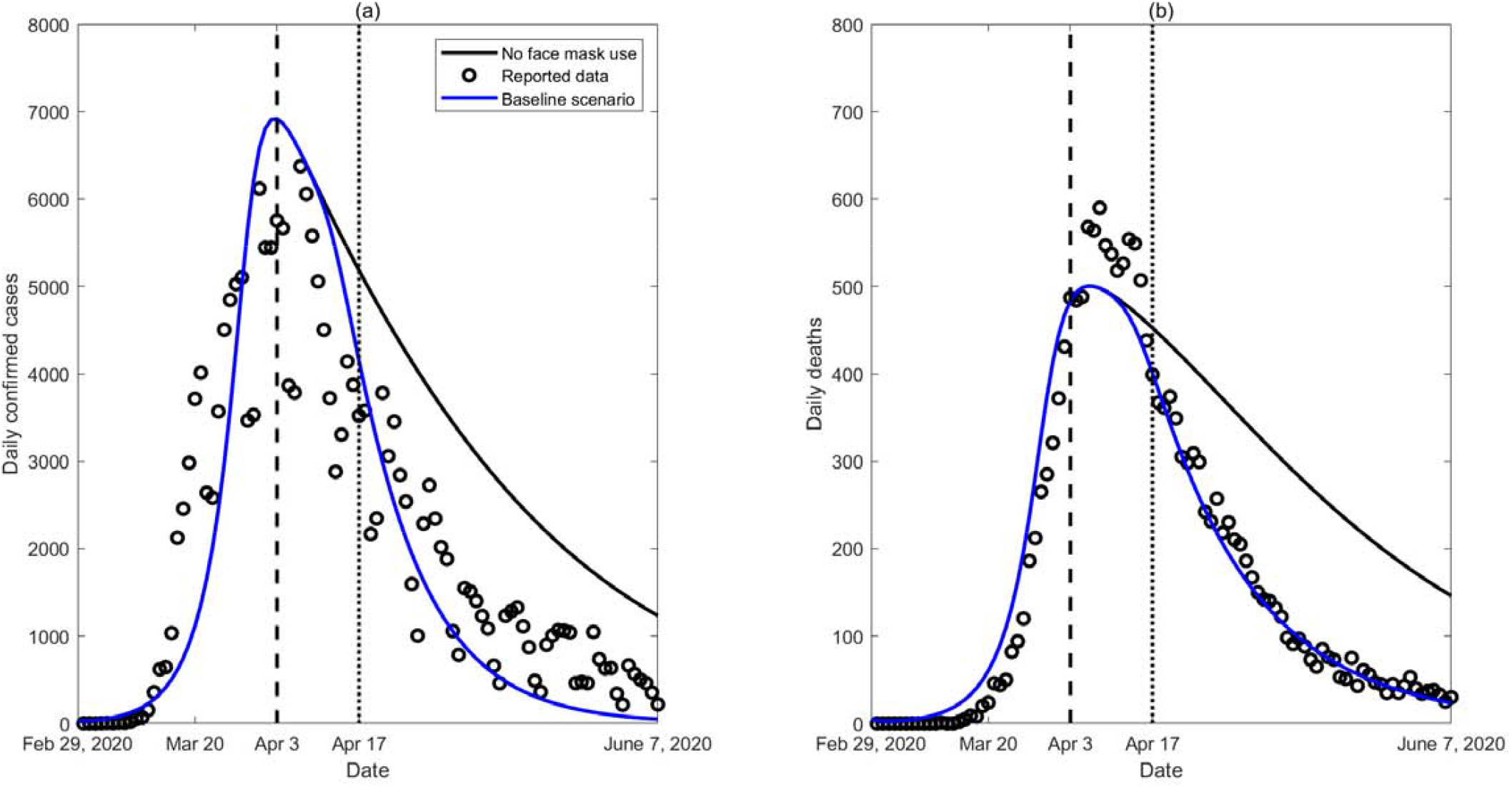
Model calibration based on reported confirmed COVID-19 cases and deaths in New York City. Two scenarios are plotted: (1) the status quo (blue lines) of Executive Order on face mask use implemented on April 17 after April 3 when the CDC began to recommend face mask use (baseline scenario); and (2) the hypothetical scenario without face mask use (solid black lines).

**Figure 3.**
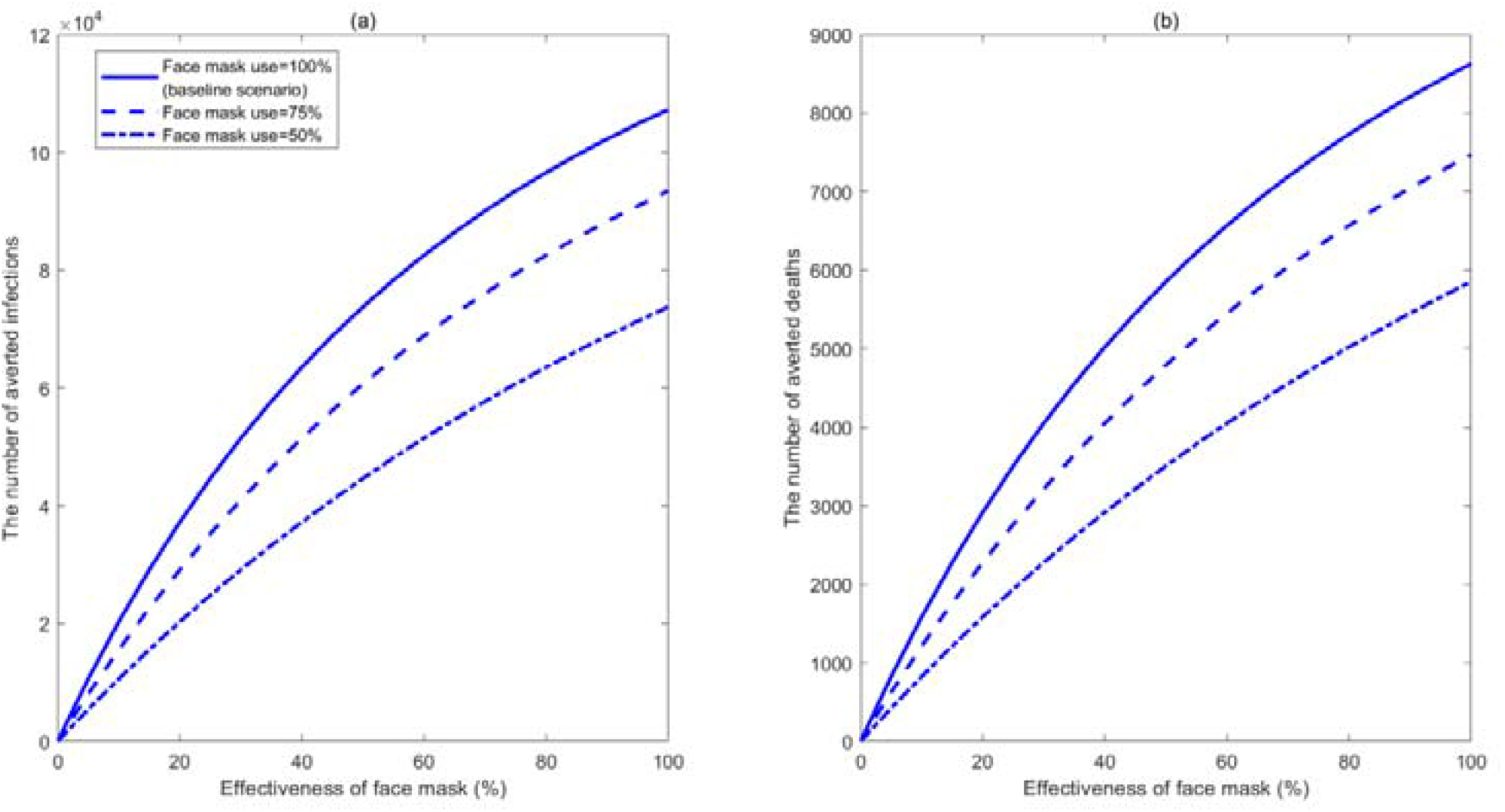
Estimated number of averted infections (a) and deaths (b) from February 29 to June 7, 2020, compared with no face mask use, for different effectivenesses and proportions of face mask use when the Executive Order on face mask use was implemented on April 17 in New York City.

### Model calibration

We calibrated the model using data on daily and cumulative confirmed infections and deaths, and estimated parameters using a nonlinear least-squares method. The unknown parameters (**Table S2**) were sampled within their ranges by the Latin hypercube sampling method and repeated 1,000 times. For every simulation, we calculated the sum of square errors between the model output and data and selected the top 10% with the least square errors to generate 95% confidence intervals. The detailed calibration procedure is provided in the **Supplementary Appendix**. All analyses and simulations were performed in MATLAB R 2019b.

### Construction of scenarios

We constructed four scenarios: (1) the status quo for which the Executive Order on face mask use was implemented on April 17 (baseline scenario); and three hypothetical scenarios, including (2) no face mask use; (3) the Executive Order was implemented one week earlier on April 10; and (4) the Executive Order was implemented two weeks earlier on April 3, the same day the CDC started recommending face mask use by the general public. We simulated the epidemic trend of infections and deaths in the above four scenarios and plotted them for scenario (1) and (2) in **Figure 2**. We also calculated the number of averted infections and deaths from February 29 to June 7 for scenarios (1), (3) and (4), compared with scenario (2).

### Sensitivity analysis

We performed sensitivity analyses on the effectiveness of face masks and proportion of face mask use and estimated how they would affect the number of averted infections and deaths.

## RESULTS

Under the status quo, the estimated number of COVID-19 infections and deaths from February 29 to June 7 were 195,617 (95% CIs: 151,580-239,653) and 17,322 (13,144-21,501), which were well consistent with reported data, as shown in **Figure 2**. In the absence of face mask use, the estimated number of infections and deaths would be 295,134 (224,459-365,809) and 25,301 (18,845-31,756), respectively **(Table 1**), indicating 99,517 (72,723-126,312) infections and 7,978 (5,692-10,265) deaths would have been averted through face mask use.

**Table 1.** The number of cumulative infections and deaths from February 29 to June 7, 2020, with the Executive Order on face mask use initiation dates of April 17, April 10, April 3 and no face mask use.

| **Simulation scenarios** | **Cumulative infections** | **Averted infections** | **Cumulative deaths** | **Averted deaths** |
| --- | --- | --- | --- | --- |
| **No face mask use** | 295,134<br>(224,459-365,809) | -- | 25,301<br>(18,845-31,756) | -- |
| **April 17 (baseline scenario)** | 195,617<br>(151,580-239,653) | 99,517<br>(72,723-126,312) | 17,322<br>(13,144-21,501) | 7,978<br>(5,692-10,265) |
| **April 10** | 183,659<br>(142,710-224,608) | 111,475<br>(81,593-141,356) | 16,284<br>(12,389-20,178) | 9,017<br>(6,446-11,589) |
| **April 3** | 166,536<br>(129,938-203,133) | 128,598<br>(94,373-162,824) | 14,786<br>(11,295-18,277) | 10,515<br>(7,540-13,489) |

If the Executive Order was implemented one week earlier on April 10, the estimated number of infections and deaths were 183,659 (142,710-224,608) and 16,284 (12,389-20,178), respectively (**Table 1**), suggesting that 111,475 (81,593-141,356) infections and 9,017 (6,446-11,589) deaths would be averted compared to no face mask use. If the Executive Order was implemented two weeks earlier on April 3, the same day as the CDC started recommending face mask use, 128,598 (94,373-162,824) infections and 10,515 (7,540-13,489) deaths would have been averted.

The sensitivity analysis (**Figure 3**) showed that when applied to the status quo, if the effectiveness of a face mask is 50%, the Executive Order would have averted 73,757 (53,663-93,850) infections and 5,852 (4,157-7,547) deaths. Even if the effectiveness of face mask was as low as 20%, the Executive Order would still avert 37,206 (26,944-47,468) infections and 2,916 (2,062-3,771) deaths. If the effectiveness of a face mask is 50% but the proportion of fack mask use decreased to 75%, the Executive Order would have averted 60,678 (44,071-77,285) infections and 4,792 (3,398-6,186) deaths. If the proportion of fack mask use further decreased to 50%, the Executive Order would have averted 44,632 (32,346-56,917) infections and 3,506 (2,481-4,531) deaths. This suggests that even with reduced effectiveness and use rate, the Executive Order on face mask use would prevent a large number of COVID-19 infections and deaths.

## DISCUSSION

This study assessed the effect of an Executive Order on face mask use on the spread of COVID-19 in NYC. We estimated that the Executive Order implemented on April 17 would avert approximately 100,000 infections and 8,000 deaths in NYC, compared with no face mask use. Findings from our study are consistent with those reported in another recent study that modelled the impact of face mask use.^14^ If the Executive Order was implemented a week earlier, it would avert 10,000 additional infections and 1,000 additional deaths. This powerful effect would come at a minimal financial cost, particularly compared to some contact reduction measures such as closing restaurants or other social venues.

Our findings suggest that increasing face mask use would significantly reduce the spread of COVID-19 in NYC. This finding is consistent with a recent study in Wuhan, China, which demonstrated that high face mask use was necessary to prevent a second major outbreak in the city after the lifting of the metropolitan-wide quarantine.^15^ Face mask use by the general public is further supported by similar practices in Singapore and Hong Kong where the governments have required their residents to wear masks in public settings and the changing position of the World Health Organization and the CDC to recommend mass population use of face masks.^7,9,16–19^ More importantly, a large proportion of transmission may occur from individuals infected by COVID-19 but showing no specific signs of symptoms, and face mask use by the general public may prevent these asymptomatic individuals from spreading the virus.^20^ Ensuring face mask use by the general public is necessary to maximize the chances of curbing the pandemic and reducing the number of COVID-19 related hospitalizations and deaths.

Widespread face mask use by the general public faces a number of challenges. First, a change in perceptions related to face mask use requires time and effort. An online survey conducted in May, 2020, showed that about 78% of survey participants wore a face mask in public and participants in those states with an Excecutive Order on face mask use were more likely to wear face masks.^21^ This is a substantial improvement from the previous unwillingness and ambivalence of using a face mask but the usage level is still lower than other countries such as South Korea (89%).^22^ Second, there is a face mask shortage in NYC and across the US in general, which has led to the prioritization of face masks for healthcare workers and those in direct contact with infected individuals. To cope with the shortage, the CDC has recommended the use of homemade cloth face coverings in public settings and areas of significant community-based transmission.^6^ Although homemade face coverings may not be as effective as face masks, it will likely still have some effectiveness in transmission prevention. Our sensitivity analysis showed that even with a 20% efficacy, the Executive Order on face mask use would still avert about 37,000 infections and 3,000 deaths in NYC. Our study has several limitations and practical simplifying assumptions. First, we assumed a homogenous use of face masks among the general population. In reality, people with symptoms are more likely to wear a face mask, whereas people with no symptoms are less likely to wear a face mask. Second, the model did not identify people who are specifically at high-risk, such as people living in a population-dense area and people who are in close contact with infected individuals. Third, we did not model COVID-19 infections and deaths across different racial/ethnic groups due to the lack of such granular data over the study period. Lastly, the effectiveness of face mask varies considerably across different types of masks. To account for parameter uncertainty, we estimated the effectiveness of face mask based on a most recent meta-analysis and performed sensitivity analyses on the key model parameters.^12^

## CONCLUSIONS

Our study indicates that the Executive Order on face mask use may play an important role in mitigating the burden of COVID-19 in NYC. The earlier the Executive Order was implemented, the more infections and deaths would be averted. This will help policymakers make more informed decisions on whether and when policies on face mask use should be implemented across the country.

## Data Availability

Data are attached in the Appendix

## Declarations

### Ethics approval and consent to participate

This study used only publicly available secondary data so ethics approval is not required.

### Consent for publication

Not applicable.

### Availability of data and materials

The COVID-19 data used in this modelling study can be found in the Supplementary Document and the NYC Department of Health and Mental Hygiene website: https://www1.nyc.gov/site/doh/covid/covid-19-main

## Authors’ contributions

Y.L., L.Z., M.S. and J.Z. conceived and designed the study. M.S. and J.Z. analyzed the data. M.S. and J.Z. carried out the analysis and performed numerical simulations. C.F., J.A.P., B.F., B.L., S.S.Y., E.C., G.L., Y.G., L.R., Y.X., G.Z. A.Z., B.C. contributed to the validation and interpretation of the results. All the authors contributed to the writing the paper, critical revision of the first draft, and approved the final manuscript for submission.

## Acknowledgements

This work was supported by the National Natural Science Foundation of China (81950410639 (LZ), 11801435 (MS), 11631012 (YX)); Outstanding Young Scholars Support Program (3111500001(LZ)); Xi’an Jiaotong University Basic Research and Profession Grant (xtr022019003(LZ), xzy032020032 (LZ)) and Xi’an Jiaotong University Young Scholar Support Grant (YX6J004(LZ)); the Bill & Melinda Gates Foundation (20200344(LZ)); China Postdoctoral Science Foundation (2018M631134, 2020T130095ZX); the Fundamental Research Funds for the Central Universities (xjh012019055, xzy032020026); Natural Science Basic Research Program of Shaanxi Province (2019JQ-187); Xi’an Special Science and Technology Projects on Prevention and Treatment of Novel Coronavirus Penumonia Emergency (20200005YX005); Zhejiang University special scientific research fund for COVID-19 prevention and control (2020XGZX056).

