## supplementary appendix for "Effects of New York’s Executive Order on Face Mask Use on COVID-19 Infections and Mortality: A Modeling Study"

This supplementary document describes in detail the mathematical model and estimation of parameters presented in the main text.

**1. Model formulation**

We proposed a dynamic compartmental model to describe the transmission of COVID-19 in New York City. The population is divided into nine compartments (**Figure S1**): susceptible individuals (S), latent individuals (E), asymptomatic infections (A), undiagnosed infections with mild/moderate (I_1_) and severe/critical symptoms (I_2_), diagnosed infections with mild/moderate (T_1_) and severe/critical symptoms (T_2_), recovered (R) and deceased (D) cases. The total population size is denoted by N, where N=S+E+A+ I_1_+ I_2_+T_1_+ T_2_ +R.

S

E

I_1_

T_1_

R

D

I_2_

T_2_

A

$$\Lambda$$

$$k_{1}\rho$$

$$k_{1}(1-\rho)$$

$$k_{2}$$

$$k_{3}$$

$$\gamma_{0}$$

$$\gamma_{0}$$

$$\alpha_{1}$$

$$\alpha_{2}$$

$$\gamma_{1}$$

$$\gamma_{2}$$

$$\mu_{1}$$

$$\mu_{2}$$

**Figure S1**. A schematic flow diagram of the transmission of COVID-19.

Susceptible individuals become infected by contacts with latent, asymptomatic and undiagnosed infectious individuals with symptoms in the public settings (e.g. public transportations, supermarkets, offices, etc) and household (home or other private settings). The total force of infection $\Lambda$ is given by the sum of forces of infections via these routes. That is,

$\Lambda= \Lambda_{pub}+\Lambda_{pri}.$	⑴

For each of route of transmission,

(a) public contacts

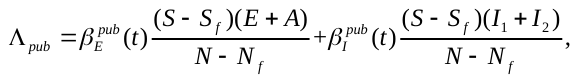
 ⑵

(b) household contacts

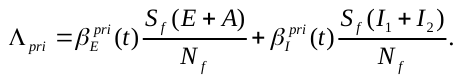
 ⑶

where

$\beta_{I}^{pub}(t)=\beta m_{1}(t)(1-\theta_{1}p_{1}(t))(1-\theta_{2}q(t)),\beta_{E}^{pub}(t)=(1-\varepsilon)\beta_{I}^{pub}(t),$	$\beta_{I}^{pri}\left( t \right)=\beta m_{2}\left( t \right)\left( 1-\theta_{1}p_{2} \right)(1-\theta_{2}q(t)),\beta_{E}^{pri}\left( t \right)=\left( 1-\varepsilon\right)\beta_{I}^{pri}\left( t \right).$	⑷

Here $\beta$ denotes the probability of transmission per contact with the infectious individuals with symptoms. We assumed that for contacts with the latent and asymptomatic individuals this probability is lower, i.e. $(1-\varepsilon)\beta$ where $0\leq\varepsilon\leq1$ denotes the reduction in per-act transmission probability. The parameters *m*_1_(*t*) and *m*_2_(*t*) represent the average number of daily person-to-person contacts in the public settings and household, *p*_1_(*t*) and *p*_2_ denote the proportion of mask usage in the public settings and household, respectively, and $\theta_{1}$ is the effectiveness of face mask/respirators in infection prevention. Let $q(t)$ and $\theta_{2}$ denote the proportion of handwashing (assumed the same in public settings and household) and the effectiveness of handwashing in infection prevention.

*Estimation of the population size and number of susceptibles for each route of transmission*

For household contacts, the overall population size (*N_f_*) is estimated as the total number of households members that are at risk of COVID-19 infection, whereas the number of susceptible households members (*S_f_*) is the difference between *N_f_* and the number of infected individuals in these households. We assumed that the number of the households at risk of infection is the same as the number of individuals infected in public settings because the probability of two or more household members being infected at the same time but at different public venues is very small. Hence, the entry of *N_f_* is$r\Lambda_{pub}$, where *r* is the average number of household members in a US family and $\Lambda_{pub}$ is the number of individuals infected in public settings as shown in eq. (2). We assumed that the infected family members become recovered after the mean period $1/\xi$. Further, the entry of susceptible household members *S_f_* is $\left( r-1 \right)\Lambda_{pub}-\Lambda_{pri}$, where $\Lambda_{pri}$ denotes the number of infected household members through household transmission as shown in eq. (3).

For public contacts, the overall population size is the number of residents (*N*) in New York City minus the overall population size (*N_f_*) in household contacts, whereas the number of susceptibles is the number of individuals free of COVID-19 infection (*S*) minus the susceptibles (*S_f_*) in household contacts.

*Modeling disease progression*

Individuals in the incubation period (*E*) progress to the infectious compartment with mild/moderate symptoms or asymptomatic compartment at a rate *k*_1_ and the probability that an individual is asymptomatic is $\rho$. Infectious individuals with mild/moderate and severe/critical symptoms are diagnosed and treated at the rates $\alpha_{1}$and $\alpha_{2}$, respectively. We assume these diagnosed individuals are isolated strictly and could not further infect others. Undiagnosed and diagnosed mild/moderate cases progress to the severe/critical stage at the rates $k_{2}$and $k_{3}$, respectively. Asymptomatic infections and undiagnosed mild/moderate cases are assumed to recover naturally at the rate $\gamma_{0}$. Diagnosed mild/moderate and severe/critical cases will recover at the rates $\gamma_{1}$ and $\gamma_{2}$, respectively. Undiagnosed and diagnosed severe/critical cases will die due to the disease at the rates $\mu_{1}$ and $\mu_{2}$, respectively. The model is described by the following system of ordinary differential equations:

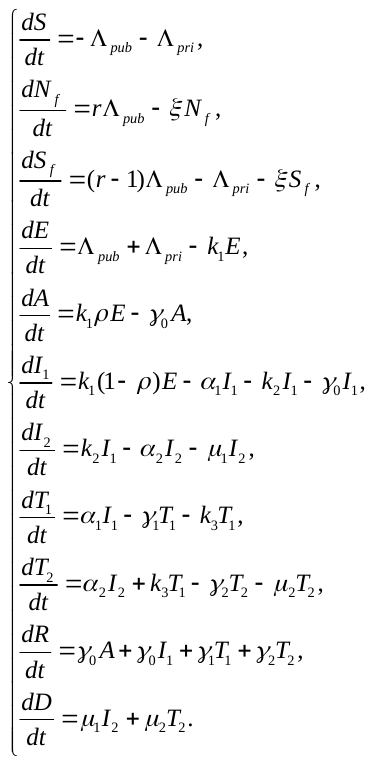
⑸

The cumulative number of deaths is tracked by the last equation of *D* in eq. (5) and the cumulative number of confirmed cases *C* is governed by the equation

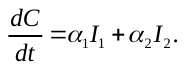
 ⑹

**2. Data and parameter estimation**

We collected the data on the number of daily and cumulative confirmed cases and deaths from February 29 to June 7, 2020 from the New York City Department of Health and Mental Hygiene (**Table S1**).^1^ The mean incubation time for COVID-19 is about 5.2 days (1/*k*_1_=5.2).^2^ The mean time from the onset of symptoms to severe/critical symptoms was 10 days (1/*k*_2_=10).^3^ The mean number of members in a US household is 3.14, so we assumed *r*=4.^4^ The average period from the onset of symptoms to diagnosis is 7 days and from the onset of symptoms to recovery for those with mild/moderate symptoms is 2 weeks in China,^5^ which indicates that the average recovery period for diagnosed mild/moderate cases is 14-7=7 days, so we assume the same average recovery period in New York City, i.e., 1$/\gamma_{1}$=7. Similarly, the average period from the onset of symptoms to recovery for those with severe/critical symptoms is 3 weeks,^6^ we have $\frac{1}{\gamma_{2}}+\frac{1}{\alpha_{2}}=21$. We assumed that the duration of recovery for infected family members is 4 weeks ($1/{\xi=28)}$.^7^ The probability of transmission per contact with latent and asymptomatic individuals is assumed to be 25% ($1-\varepsilon$=0.25) of that with infectious symptomatic individuals.^8^

The data from China^9^ and UK^10^ show that the number of daily contacts decreased by 80% and 74%, respectively after the city quarantine. Similarly, we assumed that the average number of daily contacts in the public settings $m_{1}(t)$ (**Figure S2a**) can be reduced by up to 80% in New York City in the base case,^11^ described by a decreasing logistic function${m_{1}\left( t \right)=m}_{ini}+\frac{0.2m_{ini}-m_{ini}}{1+exp(-m_{0}(t-t_{ini} ))}$, where $m_{ini}$ is the background daily contact number in the public settings before the city was put “on pause”,^12^ $m_{0}$ is the change rate of contact number, $t_{ini}$ is the time when contact number is the half of initial and minimal contact number in public settings and these three parameters will be estimated by model fitting. Home confinement led to double contact rate than the pre-quarantine level.^13^ We assumed that the average number of daily contacts in a household (**Figure S2a**) increased from 4 to 8, described by an increasing logistic function $m_{2}\left( t \right)=4+\frac{8-4}{1+exp(-m_{0}(t-t_{ini}))}$.

The proportion of handwashing in the US was assumed to increase during the outbreak.^14^ We assumed a logistic growth for this percentage (**Figure S2b**), i.e. ${q\left( t \right)=q}_{ini}+\frac{\bar{q}-q_{ini}}{1+exp(-m_{0}(t-t_{ini}))}$ where $q_{ini}$ is the base proportion of handwashing before the outbreak, and $\bar{q}$ is the maximum proportion of handwashing during the outbreak. A survey on handwashing culture from 63 countries showed that about 77% of people in US had a habit of automatic handwashing in routine life,^14^ so we chose $p_{ini}=77\%$ as the base value of the proportion of handwashing in the absence of COVID-19. Another online survey in US showed that 88.27-99% of people washed hands during the epidemic,^15^ so we assumed $\bar{q}=95\%$ as the maximum proportion of handwashing in New York City. The effectiveness of handwashing in preventing infection ($\theta_{2}$) is chosen to be 42% (50-95%), based on a review against respiratory infections.^16^

US CDC recommended face mask use in April 3, and after two weeks NYC implemented the Executive Order on face mask use that required all residents over age 2 must wear masks or face coverings when they're in public and social distancing isn't possible.^17^ We assumed its use coverage increased from almost 0 on April 3 to 100% on April 17. We assumed a logistic growth for this percentage (**Figure S2c**), i.e. ${p_{1}\left( t \right)=p}_{ini}+\frac{\bar{p}-p_{ini}}{1+exp(-(t-t_{p}))}$ where $p_{ini}=0$ is the base proportion of facial mask usage in the public settings before CDC recommendation, $\bar{p}=100\%$ is the maximum proportion of facial mask usage in the public settings after the Executive Order on face mask use, and $t_{p}=42$ is the time when the proportion of face mask usage is the half of base and maximum proportion (Apr 3 minus Feb 29 plus one week). The coverage ratio of face mask use in private settings $p_{2}$ is set to zero.^18^ The effectiveness of face mask in preventing infection ($\theta_{1}$) is chosen to be 85% (66-93%), based on a meta-analysis against COVID-19.^19^ We varied the effectiveness of face mask from 0 to 100% in sensitivity analysis (**Figure 3**).

The total population size in NYC is 8,398,748. The initial values of the disease states are given by T_1_(0)=0, T_2_(0)=0, R(0)=0, D(0)=0, N(0)= 8,398,748, $N_{f}\left( 0 \right)=rC\left( 0 \right)=4$, and $S_{f}\left( 0 \right)=(r-1)C\left( 0 \right)=3$. We left E(0),A(0), I_1_(0), I_2_(0) to be estimated by the fitting.

We calibrated the model by the daily and cumulative confirmed cases and deaths from February 29 to June 7, 2020, when NYC was not reopened, by using a nonlinear least-squares method (**Figure 2, Figure S3**). The unknown parameters (**Table S2**) were sampled within their ranges by the Latin hypercube sampling method and repeated 1000 times. For every simulation, we calculated the sum of square errors between the model output and data, and selected the top 10% with the least square errors to generate 95% confidence intervals.

Based on the estimated parameters, we simulate the epidemic trend of daily confirmed cases and deaths (**Figure 2**). We also calculate the number of averted infections and deaths during February 29 to June 7, 2020, compared with no face mask use, for the Executive Order on face mask use initiation dates of April 17, April 10, April 3 as shown in**Table 1.**

**Table S1**. Reported daily and cumulative confirmed cases and deaths data in New York City from the NYC Department of Health and Mental Hygiene^1^

| **Date** | **New COVID-19 cases** | **New deaths** | **Cumulative confirmed cases** | **Cumulative deaths** |
| --- | --- | --- | --- | --- |
| 2020-2-29 | 1 | 0 | 1 | 0 |
| 2020-3-1 | 1 | 0 | 2 | 0 |
| 2020-3-2 | 0 | 0 | 2 | 0 |
| 2020-3-3 | 2 | 0 | 4 | 0 |
| 2020-3-4 | 5 | 0 | 9 | 0 |
| 2020-3-5 | 3 | 0 | 12 | 0 |
| 2020-3-6 | 8 | 0 | 20 | 0 |
| 2020-3-7 | 7 | 0 | 27 | 0 |
| 2020-3-8 | 21 | 0 | 48 | 0 |
| 2020-3-9 | 58 | 0 | 106 | 0 |
| 2020-3-10 | 70 | 0 | 176 | 0 |
| 2020-3-11 | 154 | 1 | 330 | 1 |
| 2020-3-12 | 357 | 0 | 687 | 1 |
| 2020-3-13 | 620 | 0 | 1307 | 1 |
| 2020-3-14 | 644 | 2 | 1951 | 3 |
| 2020-3-15 | 1034 | 5 | 2985 | 8 |
| 2020-3-16 | 2123 | 9 | 5108 | 17 |
| 2020-3-17 | 2455 | 8 | 7563 | 25 |
| 2020-3-18 | 2981 | 20 | 10544 | 45 |
| 2020-3-19 | 3714 | 24 | 14258 | 69 |
| 2020-3-20 | 4013 | 46 | 18271 | 115 |
| 2020-3-21 | 2639 | 44 | 20910 | 159 |
| 2020-3-22 | 2581 | 50 | 23491 | 209 |
| 2020-3-23 | 3568 | 82 | 27059 | 291 |
| 2020-3-24 | 4504 | 94 | 31563 | 385 |
| 2020-3-25 | 4846 | 120 | 36409 | 505 |
| 2020-3-26 | 5026 | 186 | 41435 | 691 |
| 2020-3-27 | 5103 | 212 | 46538 | 903 |
| 2020-3-28 | 3464 | 265 | 50002 | 1168 |
| 2020-3-29 | 3530 | 285 | 53532 | 1453 |
| 2020-3-30 | 6120 | 321 | 59652 | 1774 |
| 2020-3-31 | 5444 | 372 | 65096 | 2146 |
| 2020-4-1 | 5444 | 431 | 70540 | 2577 |
| 2020-4-2 | 5755 | 487 | 76295 | 3064 |
| 2020-4-3 | 5667 | 484 | 81962 | 3548 |
| 2020-4-4 | 3865 | 488 | 85827 | 4036 |
| 2020-4-5 | 3784 | 568 | 89611 | 4604 |
| 2020-4-6 | 6375 | 564 | 95986 | 5168 |
| 2020-4-7 | 6056 | 590 | 102042 | 5758 |
| 2020-4-8 | 5579 | 547 | 107621 | 6305 |
| 2020-4-9 | 5057 | 537 | 112678 | 6842 |
| 2020-4-10 | 4503 | 518 | 117181 | 7360 |
| 2020-4-11 | 3722 | 526 | 120903 | 7886 |
| 2020-4-12 | 2879 | 554 | 123782 | 8440 |
| 2020-4-13 | 3304 | 549 | 127086 | 8989 |
| 2020-4-14 | 4139 | 507 | 131225 | 9496 |
| 2020-4-15 | 3874 | 438 | 135099 | 9934 |
| 2020-4-16 | 3521 | 399 | 138620 | 10333 |
| 2020-4-17 | 3577 | 367 | 142197 | 10700 |
| 2020-4-18 | 2166 | 361 | 144363 | 11061 |
| 2020-4-19 | 2344 | 374 | 146707 | 11435 |
| 2020-4-20 | 3781 | 349 | 150488 | 11784 |
| 2020-4-21 | 3059 | 305 | 153547 | 12089 |
| 2020-4-22 | 3452 | 298 | 156999 | 12387 |
| 2020-4-23 | 2841 | 309 | 159840 | 12696 |
| 2020-4-24 | 2538 | 299 | 162378 | 12995 |
| 2020-4-25 | 1595 | 242 | 163973 | 13237 |
| 2020-4-26 | 1004 | 231 | 164977 | 13468 |
| 2020-4-27 | 2285 | 257 | 167262 | 13725 |
| 2020-4-28 | 2725 | 218 | 169987 | 13943 |
| 2020-4-29 | 2342 | 230 | 172329 | 14173 |
| 2020-4-30 | 2016 | 210 | 174345 | 14383 |
| 2020-5-1 | 1880 | 205 | 176225 | 14588 |
| 2020-5-2 | 1057 | 186 | 177282 | 14774 |
| 2020-5-3 | 785 | 167 | 178067 | 14941 |
| 2020-5-4 | 1547 | 150 | 179614 | 15091 |
| 2020-5-5 | 1510 | 142 | 181124 | 15233 |
| 2020-5-6 | 1401 | 140 | 182525 | 15373 |
| 2020-5-7 | 1229 | 132 | 183754 | 15505 |
| 2020-5-8 | 1085 | 122 | 184839 | 15627 |
| 2020-5-9 | 661 | 98 | 185500 | 15725 |
| 2020-5-10 | 461 | 91 | 185961 | 15816 |
| 2020-5-11 | 1232 | 97 | 187193 | 15913 |
| 2020-5-12 | 1284 | 88 | 188477 | 16001 |
| 2020-5-13 | 1326 | 73 | 189803 | 16074 |
| 2020-5-14 | 1111 | 65 | 190914 | 16139 |
| 2020-5-15 | 871 | 85 | 191785 | 16224 |
| 2020-5-16 | 489 | 76 | 192274 | 16300 |
| 2020-5-17 | 361 | 73 | 192635 | 16373 |
| 2020-5-18 | 901 | 53 | 193536 | 16426 |
| 2020-5-19 | 1006 | 51 | 194542 | 16477 |
| 2020-5-20 | 1068 | 75 | 195610 | 16552 |
| 2020-5-21 | 1063 | 43 | 196673 | 16595 |
| 2020-5-22 | 1041 | 61 | 197714 | 16656 |
| 2020-5-23 | 462 | 56 | 198176 | 16712 |
| 2020-5-24 | 475 | 47 | 198651 | 16759 |
| 2020-5-25 | 462 | 45 | 199113 | 16804 |
| 2020-5-26 | 1047 | 35 | 200160 | 16839 |
| 2020-5-27 | 736 | 45 | 200896 | 16884 |
| 2020-5-28 | 631 | 35 | 201527 | 16919 |
| 2020-5-29 | 635 | 43 | 202162 | 16962 |
| 2020-5-30 | 341 | 53 | 202503 | 17015 |
| 2020-5-31 | 215 | 40 | 202718 | 17055 |
| 2020-6-1 | 664 | 34 | 203382 | 17089 |
| 2020-6-2 | 569 | 37 | 203951 | 17126 |
| 2020-6-3 | 503 | 38 | 204454 | 17164 |
| 2020-6-4 | 462 | 33 | 204916 | 17197 |
| 2020-6-5 | 354 | 25 | 205270 | 17222 |
| 2020-6-6 | 219 | 30 | 205489 | 17252 |
| 2020-6-7 | 181 | 28 | 205670 | 17280 |

**Table S2**. The values of parameters based on references or estimated by nonlinear least-squares (NLS) method.

| Parameter | Description | Range or 95% CI from NLS | Source |
| --- | --- | --- | --- |
| 1/*k*_1_ | The mean incubation time (days) | 5.2 (4.1-7.0) | ^2^ |
| 1$/k_{2}$ | The mean time from symptoms onset to natural recovery (days) | 10 | ^3^ |
| $r$ | The mean number of members in a family | 4 | ^4^ |
| 1$/\gamma_{1}$ | The average recovery period for diagnosed mild/moderate cases (days) | 7 | ^5,6^ |
| 1/$\gamma_{0}$ | The mean time for natural recovery (days) | 6.9980 (6.8881-7.1113) | NLS |
| 1$/\alpha_{1}$ | The average period from symptoms onset to diagnose for mild/moderate cases (days) | 4.9998 (4.7309-5.3012) | NLS |
| 1$/\alpha_{2}$ | The average diagnose period for severe/critical cases (days) | 2.0973 (2.0018-2.2023) | NLS |
| 1$/\gamma_{2}$ | The average recovery period for diagnosed severe/critical cases | $21-1/\alpha_{2}$ | ^6^ |
| $1/\xi$ | The mean recovery period for infected family members (days) | 28 | Assumed |
| E(0) | The initial value of latent individuals | 491.1294 (488.8572-493.4017) | NLS |
| A(0) | The initial value of asymptomatic individuals | 99.9997 (98.8617-101.1376) | NLS |
| I_1_(0) | The initial value of undiagnosed mild/moderate cases | 10.2234 (9.0856-11.3611) | NLS |
| I_2_(0) | The initial value of undiagnosed severe/critical individuals | 50.0007 (48.8631-51.1384) | NLS |
| $\beta$ | The per-act transmission probability in contact with infected individuals with symptoms | 0.0379 (0.0376-0.0382) | NLS |
| $\varepsilon$ | The reduction in per-act transmission probability if infection is in latent and asymptomatic stage | 75% | ^8^ |
| $\rho$ | The probability that an individual is asymptomatic | 0.3841 (0.3386-0.4296) | NLS |
| $k_{3}$ | The progression rate from diagnosed mild/moderate stage to diagnosed severe/critical stage | 0.0040 (0.0006-0.0074)$\times k_{2}$ | NLS |
| $m_{1}(t)$ | The average number of daily contacts in the public settings | $m_{ini}+\frac{0.2m_{ini}-m_{ini}}{1+\exp(-m_{0}(t-t_{ini}))}$ | ^12,20^ |
| $m_{ini}$ | Base daily contact number in the public settings | 26.7629 (26.6491-26.8767) | NLS |
| $m_{0}$ | Change rate of daily contact number | 0.8742 (0.8515-0.8970) | NLS |
| $t_{ini}$ | The time when the contact number is half of initial and minimal contact number in public settings | 27.9999 (27.7727-28.2272) | NLS |
| $m_{2}(t)$ | The average number of daily contacts in the households | $4+\frac{12-4}{1+\exp(-m_{0}(t-t_{ini}))}$ | ^13^ |
| $q(t)$ | The usage percentage of handwashing | $q_{ini}+\frac{\bar{q}-q_{ini}}{1+\exp(-m_{0}(t-t_{ini}))}$ | ^14^ |
| $q_{ini}$ | Base percentage of handwashing before the epidemic | 77% | ^14^ |
| $\bar{q}$ | Maximal percentage of handwashing during the epidemic | 95% | ^15^ |
| $\theta_{2}$ | The effectiveness of handwashing in preventing infection | 0.42 (0.1-0.95) | ^16^ |
| $p_{1}(t)$ | The usage percentage of face mask in the public settings | $p_{ini}+\frac{\bar{p}-p_{ini}}{1+\exp(-(t-t_{p}))}$ | ^21^ |
| $p_{ini}$ | Base percentage of face mask usage in the public settings before CDC recommendation | 0% | Asumed |
| $\bar{p}$ | Percentage of face mask usage in the public settings after the Executive Order on face mask use | 100% | Assumed |
| $t_{p}$ | The time when face mask usage in the public settings is half of the maximal face mask usage rate | 42 | Assumed |
| $p_{2}$ | The usage percentage of mask in the households | 0% | ^21^ |
| $\theta_{1}$ | The effectiveness of mask in preventing infection | 0.85 (0.66-0.93) | ^19^ |
| $\mu_{1}$ | Disease-induced death rate of undiagnosed severe/critical cases | 0.1000 (0.0886-0.1114) | NLS |
| $\mu_{2}$ | Disease-induced death rate of diagnosed severe/critical cases | ${0.0524 (0.0518-0.0530)\times\mu}_{1}$ | NLS |

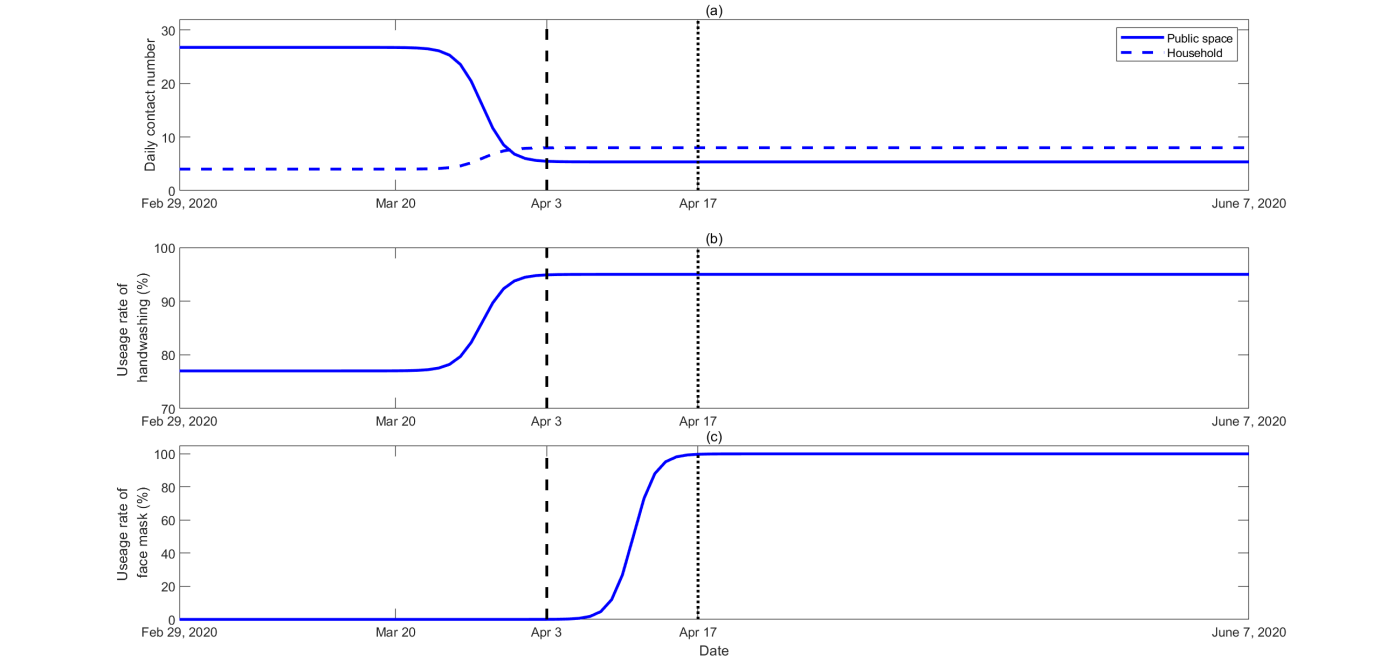

**Figure S2**. (a) The average number of daily contacts in the public settings ${m_{1}\left( t \right)=m}_{ini}+\frac{0.2m_{ini}-m_{ini}}{1+\exp(-m_{0}(t-t_{ini}))}=26.7629+\frac{0.2\times26.7629-26.7629}{1+\exp(-0.8742(t-27.9999))}$ over time (blue solid line) and in the households $m_{2}\left( t \right)=4+\frac{12-4}{1+\exp(-0.8742(t-27.9999))}$ over time (blue dashed line). (b) The percentage of handwashing ${q\left( t \right)=q}_{ini}+\frac{\bar{q}-q_{ini}}{1+\exp(-m_{0}(t-t_{ini}))}=77\%+\frac{95\%-77\%}{1+\exp(-0.8742(t-27.9999))}$ over time. (c) The percentage of face mask use ${p_{1}\left( t \right)=p}_{ini}+\frac{\bar{p}-p_{ini}}{1+\exp(-(t-t_{p}))}=0\%+\frac{100\%-0\%}{1+\exp(-(t-42))}$ over time. CDC began to recommend face mask use on April 3^rd^ (black dashed lines) and NYC implemented the Executive Order on face mask use on April 17^th^ (black dot lines).

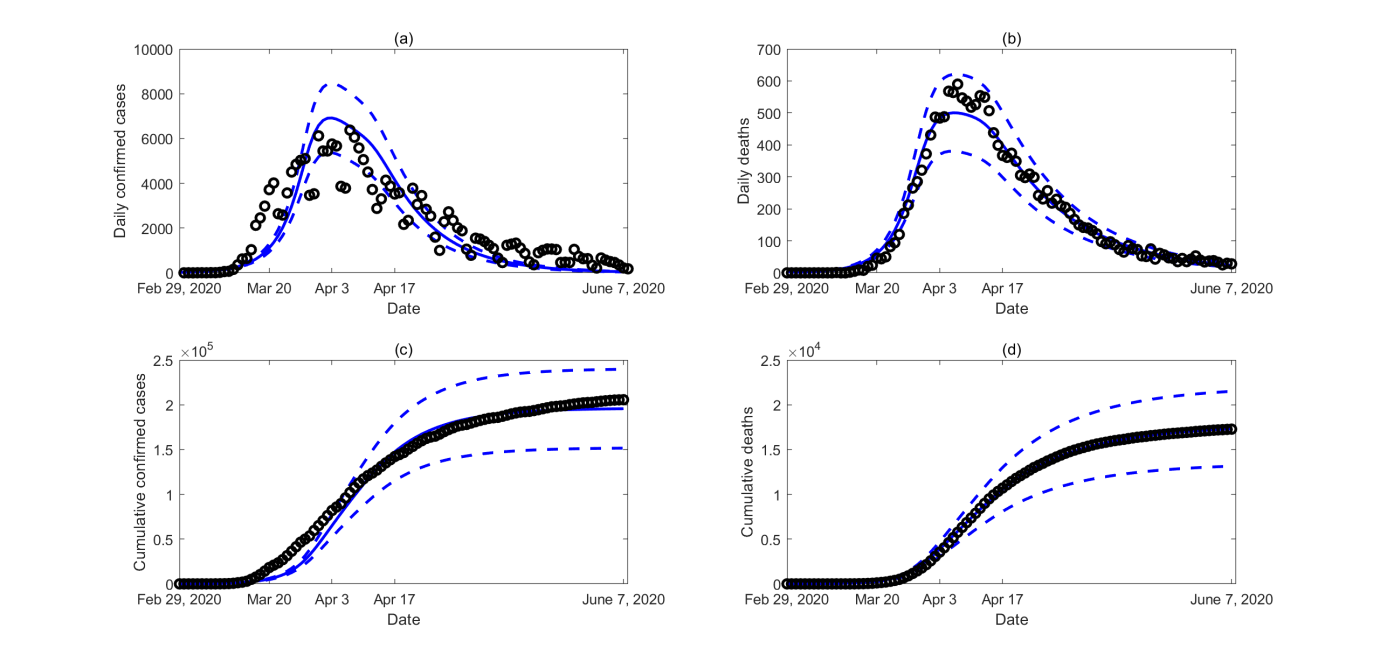

**Figure S3**. Model calibration by the number of daily and cumulative confirmed cases and deaths. Dashed lines denote 95% confidence intervals.
